# Atherosclerosis biomarkers via mitochondrial permeability transition necrosis gene analysis

**DOI:** 10.1101/2025.09.09.25335456

**Authors:** Fei-Niu, Su-Ying Wu, Yang Liu, Jing-Wen Zhang, Yong-Hao Jiang, Ying-Xuan Shi

## Abstract

**Background:** Atherosclerosis (AS) is a major cause for cardiovascular disease, and mitochondrial permeability transition driven necrosis (MPTDN) is often associated with cardiovascular disease. To investigate the role of MPTDN-associated genes (MPTDNGs) in AS is the aim of this study.

**Methods:** GSE100927 and GSE66360 datasets, along with MPTDNGs were included in this study. Employing differential expression analysis, weighted gene co-expression network analysis (WGCNA), machine learning algorithms, and the integration of receiver operating characteristic (ROC) analysis with expression profiling, this study identified biomarkers. Subsequent analyses included functional analysis and immune infiltration analysis, the prediction of targeted drugs, and the molecular docking. Eventually, the expression of biomarkers was clinically validated using reverse transcription-quantitative polymerase chain reaction (RT-qPCR).

**Results:** Colony stimulating factor 1 receptor (CSF1R), tumor necrosis factor (TNF) and integrin alpha M (ITGAM) were identified as biomarkers, all exhibiting up-regulated expression in AS samples. The ROC analysis showed that their Area Under Curve (AUC) values were all greater than 0.9, which indicated that they were good at distinguishing between AS and control samples. Gene set enrichment analysis (GSEA) enrichment results showed that biomarkers positively regulated lysosome, leishmania infection and so on. Interestingly, immune infiltration analysis showed that both differential immune cells (CD4 memory resting T cells and M0 Macrophages) and differential immune gene sets (naturalKiller cell cytotoxicity and transforming growth factor-b (TGFb) family member receptor) were strongly correlated with ITGAM.The drugs were predicted, and the molecular docking illustrated that a robust binding interaction was found between CSF1R and EDICOTINIB, with a binding energy of - 8.93 kcal/mol. Lastly, the expression validation results obtained through RT-qPCR demonstrated that CSF1R exhibited up-regulation in AS samples when compared to normal samples, while TNF and ITGAM showed less significant differences between AS and normal.

**Conclusion:** CSF1R, TNF and ITGAM were identified as biomarkers in AS, which provided a basis and reference for the study of MPTDNGs in AS.

## 1. Introduction

Atherosclerosis (AS), a primary cause of cardiovascular diseases, is a chronic inflammatory condition characterized by the accumulation of cholesterol-laden low-density lipoprotein particles in the arterial walls[1, 2]. Although AS progresses slowly and remains asymptomatic for decades, the rupture of a plaque can pose a lethal threat, leading to ischemic stroke and myocardial infarction[3]. The stability of atherosclerotic plaques is a principal factor in the occurrence of asymptomatic strokes and heightens the risk for ischemic strokes and acute coronary syndromes[4]. Unstable plaques are characterized by a large necrotic core, a thin fibrous cap, and spotty calcification, accompanied by an exacerbation of inflammation[5]. The early identification of unstable plaques, which are more prone to rupture and can lead to localized thrombosis or embolism, carries significant clinical importance for the prevention of rupture or erosion[4]. Coronary heart disease, caused by AS, is prevalent globally, with most Asian countries facing significant challenges due to this condition. Recent research reports indicate that the mortality rate per 100,000 adults ranges from 103 to 366 individuals[6]. Over the past decade, there has been a notable upward trend in the burden associated with AS, particularly in East Asia where an alarming increase of 117.2% in total deaths and 115.3% in disability-adjusted life years has been observed[6]. Another study has shown that in high-income countries, since the mid-20th century, there has been a sharp decline in the incidence and mortality rates of ischemic heart disease and ischemic stroke caused by AS[7]. The standard for treating AS involves targeting modifiable risk factors to prevent cardiovascular events and restoring arterial blood flow through percutaneous or surgical revascularization[8, 9]. However, the long-term consequences of AS remain a leading cause of death in both developed and developing nations, presenting many challenges that need to be addressed[10]. Therefore, exploring novel biomarkers and underlying mechanisms aids in the comprehension of the pathogenesis and molecular rationale of disease, offering targets for the early clinical diagnosis and treatment of AS.

The mitochondria serve as the hub for cellular respiration and as regulators at the center of cell metabolism, participating in a multitude of cellular processes. Mitochondrial permeability transition (MPT) is a phenomenon that induces transient fluxes of low molecular weight solutes across the inner mitochondrial membrane. MPT-driven necrosis (MPTDN) represents a form of cell dissolution, precipitated by disturbances in the intracellular microenvironment, such as severe oxidative stress and intracellular Ca2^+^ overload, and is also contingent upon the presence of cyclophilin D[11, 12]. This process is initiated by the opening of a multi-protein channel that has yet to be clearly characterized, commonly referred to as the permeability transition pore complex-and is negatively regulated by peptidylprolyl isomerase [13]. Under certain circumstances, an excess of mitochondrial Ca2^+^ in conjunction with an overproduction of reactive oxygen species can lead to an increased conductivity of the permeability transition pore complex, thereby driving MPT as a form of necrosis[14]. It is also involved in various cellular states and is often associated with cardiovascular diseases, cancer, and neuropathies, attracting widespread interest. In tumors, dysfunctions of mitochondrial activities, particularly MPT, can lead to the depletion of mitochondrial DNA and RNA, thereby hindering the function of the mitochondrial respiratory chain and ultimately inducing cell death in tumor cells[15–19]. In cardiovascular diseases, MPTDN is closely associated with myocardial ischemia and myocardial infarction[20, 21]. AS is a leading cause of cardiovascular diseases, yet the role of MPTDN in AS remains unclear.

Therefore, this study aimed to explore the biomarkers of MPTDN during the AS process through bioinformatics methods and to analyze the relationship between these biomarkers and immunity. This provided a reference for understanding the mechanisms of MPTDN in AS and formed the basis for its therapeutic intervention.

## Materials and methods

### 2.1 Data source

The Gene Expression Omnibus (GEO) database was used for all transcriptomic data used in this study (https://www.ncbi.nlm.nih.gov/geo/). In detail, the GSE100927 dataset had blood samples from 69 AS patients and 35 controls as a training set, and GSE66360 as a validation set contained blood samples from 49 AS patients and 50 controls. Among the samples, atherosclerotic carotid artery, atherosclerotic femoral artery and atherosclerotic infra-popliteal artery were defined as AS disease samples. In addition, 39 mitochondrial permeability transition-driven necrosis-associated genes (MPTDNGs) were derived from published literature[13].

### 2.2 Differential expression analysis and weighted correlation network analysis (WGCNA)

The R package limma (version 3.54.1)[22] was used to obtain differentially expressed genes (DEGs) between the AS and control samples in the GSE100927 (adj.P.Val< 0.05 & log_2_|FC|> 1), and R package ggplot2 (version 3.3.6)[23] and ComplexHeatmap (version 2.14.0)[24] were used to draw volcano plot and heat map respectively to intuitively reflect the DEGs. Meanwhile, in order to obtain the genes related to MPTDNGs, we calculated the scores of MPTDNGS in AS and control samples by single-sample gene set enrichment analysis (ssGSEA) algorithm included in the R package gene set variation analysis (GSVA) (version 1.42.0)[25] and compared the differences between them. The MPTDNGs score was used as a trait for analysis by R package WGCNA (version 1.71)[26]. Firstly, a hierarchical clustering tree was constructed for all samples in the GSE100927 to determine whether there were outliers. Next, our choice of the most appropriate soft threshold (β) of the adjacency function was in accordance with the criteria of scale-free networks. Then, the minimum number of genes for each gene module was set to 50 genes according to the hybrid dynamic cut tree standard, and mergeCutHeight was set to 0.3 for merging to obtain the gene module. In the end, the module with the strongest correlation with the characteristic was chosen as the key module for analysing the following, and the genes it contained were called key module genes.

### 2.3 Enrichment analysis and construction of Protein-Protein Interaction (PPI) network

By intersection of DEGs and key module genes, differentially expressed MPTDNGs (DE-MPTDNGs) were obtained. The main functions and pathways of genes can be understood through enrichment analysis. Here, R package clusterProfiler (version 4.6.2)[27] has been used to perform Gene Ontology (GO) and Kyoto Encyclopedia of Genes and Genomes (KEGG) enrichment analyses on DE-MPTDNGs (adjust.p < 0.05). In order to understand the interaction of DE-MPTDNGs at the protein level, the PPI network was constructed by STRING database (https://string-db.org/) (interaction score= 0.4). Then the analysis results were imported into Cytoscape software (version 3.9.0)[28], and the subnetwork was analysed using the CytoHubba plug-in. The genes of Maximal Clique Centrality (MMC) Top10 and the genes of Degree Top10 were intersected to obtain candidate genes.

### 2.4 Machine learning screening for feature genes

On the basis of the candidate genes, the feature genes were then further screened by means of machine algorithms in order to obtain the characteristic genes. First, the R package glmnet was used to construct a logistic regression model using the Least Absolute Shrinkage and Selection Operator (LASSO) (version 4.1-4)[29]to get the graph of feature genes coefficient and the error plot of cross-validation. The second one was Support vector machines-Recursive feature elimination (SVM-RFE) analysis by R package e1071 (version 1.7-13)[30]. In both methods set nfolds = 10 for cross-validation. Finally, the genes obtained from the above two machine algorithms were taken as intersection to determine the feature genes. For the feature genes, receiver operating characteristic (ROC) curve analysis was performed to determine their ability to discriminate between AS and control samples in the GSE100927 and GSE66360, and the Wilcoxon rank sum test was carried out to compare the feature genes in the GSE100927 and GSE66360 between the AS and control groups (P value < 0.05). The feature genes with significant differences and consistent trends in both datasets were used as biomarkers in this study.

### 2.5 Gene Set Enrichment Analysis (GSEA)

According to the median expression of biomarkers in the GSE100927, the samples were divided into low and high expression groups, and the difference analysis was carried out. Then, according to the order of genes and their corresponding fold changes, c2.cp.kegg.v7.4.1.symbols.gmt has been downloaded from the Molecular Signatures Database (MSigDB) (https://www.gsea-msigdb.org/gsea/msigdb) as the background gene set. The R package clusterProfiler was used to perform the GSEA (version 4.6.2) [27], and multiple test correction was performed using the FDR method. adjust.p < 0.05 was considered to be a significant enrichment result. Finally, the top 5 pathways of the enrichment results were visualized using the enrichplot (v 1.18.0) package[31], ordered by adjust.p values.

### 2.6 Immunological microlandscape analysis

The immune microenvironment can influence cell growth and development. In the GSE100927, for screening samples, P value < 0.05 was set, and the remaining samples were analyzed by using the LM22 immune dataset in the CIBERSORT immune cell infiltration frequency analysis algorithm of 22 immune cells in all samples. In addition, the immune response gene set was downloaded through the Immunology Database and Analysis Portal (ImmPort) (https://ngdc.cncb.ac.cn/databasecommons/database/id/4959), and the GSVA score of the different samples was calculated by means of the ssGSEA function in the R package GSVA (version 1.42.0)[32]. Subsequently, The wilcox.test function in the R package stats (version 4.2.2)[33]was used for the Wilcoxon rank sum test to obtain differential immune cells and differential immune gene sets (p< 0.05), with the results visualized in a box plot by RColorBrewer (v 3.4.1) package. Finally, the correlation between biomarkers and differential immune cells, between the different immune cells and differential immune gene sets was analysed using Spearman correlation analysis (|R| > 0.4 & P< 0.05).

### 2.7 Construction of molecular regulatory network and drug prediction of biomarkers

It is crucial to understand the molecular regulation mechanism of biomarkers in AS. First, MicroRNA Target Prediction Database (miRDB) (http://mirdb.org) and NetworkAnalyst database (https://www.networkanalyst.ca/) were used to predict miRNAs and transcription factors (TFs) that regulate biomarkers, respectively. Then, the StarBase database (https://rnasysu.com/encori/) was used to predict lncRNAs corresponding to the above miRNAs. In order to screen potential drugs targeting AS, drugs that interacted with biomarkers were predicted in Drug Gene Interaction database (DGldb) (https://www.dgidb.org/). Finally, the above relationship pairs were integrated to visualize ceRNA, TF-mRNA-miRNA and biomarker-drug networks through Cytoscape software (version 3.9.0)[28].

### 2.8 Molecular docking

Molecular docking is helpful to study the interaction between biomarker and drug. First, the crystal structures corresponding to each biomarker were downloaded from the Protein Data Bank (PDB) (https://www.rcsb.org/). Then the 3D structure of the drug with the highest Query score and Interacton Score was downloaded from the PubChem database (https://pubchem.ncbi.nlm.nih.gov/). Finally, Molecular docking was performed using AutoDock Vina and results visualised in pymol (version 2.1)[34].Lower binding energy typically indicates a stronger interaction and higher binding affinity, with a threshold of ≤-5 kcal/mol generally regarded as indicative of favorable binding affinity, and a threshold of ≤-7 kcal/mol considered indicative of stable binding affinity.

### 2.9 RNA isolation and quantitative real-time polymerase chain reaction (qRT-PCR)

A total of five pairs of control and AS samples were obtained from the clinic in the Affiliated Hospital of Shandong University of Traditional Chinese Medicine. All participants were given informed consent. The study had the approval of the Affiliated Hospital of Shandong University of Traditional Chinese Medicine ethics committee (2023)(071)-KY.

All patients had signed an informed consent form. Total RNA of 10 samples were extracted using TRIzol (Ambion, Austin, USA) according to the manufacturer’s guidance. Reverse transcription of total RNA to cDNA was carried out by using SureScript-First-strand-cDNA-synthesis-kit (Servicebio, Wuhan, China) based on the manufacturer’s instructions. RT-qPCR was performed utilizing the CFX Connect Thermal Cycler (Bio-Rad, USA). The primer sequences for PCR were shown in **Supplementary Table 1**. GAPDH was as an internal reference gene. The 2^−ΔΔCt^ method was utilized to calculate the expression of biomarkers[35].

**Supplementary Table 1:** Sequence information of primers

### 2.10 Statistical analysis

All analyses were executed in R software (v 4.2.2).All P values of statistical results were based on Wilcoxon rank sum test, and statistical significance was defined as P value < 0.05.

## 3. Results

### 3.1 Identifying of DEGs and key module genes

Differential expression analysis showed that there were 409 DEGs between AS and control groups, of these, 107 were up-regulated and 302 were down-regulated **(Figure 1A-B)**. In addition, the MPTDNGs score showed that significantly higher in the AS group than in the control group **(Figure 1C)**. After that, we performed WGCAN with MPTDNGs score as a trait to obtain key module genes. The hierarchical clustering tree showed that there was no outlier sample **(Figure 1D)**. When β= 8 and R^2^= 0.9, the co-expression network was close to scale-free distribution **(Figure 1E)**.Then, using the dynamic splicing algorithm, 13 gene modules were obtained **(Figure 1F)**, of which the purple module was found to correlate the most with the MPTDNGS score (Cor= 0.87 & p< 0.05) **(Figure 1G)**. Thus, it was used as the key module, which contained 4904 key module genes.

**Figure 1:**
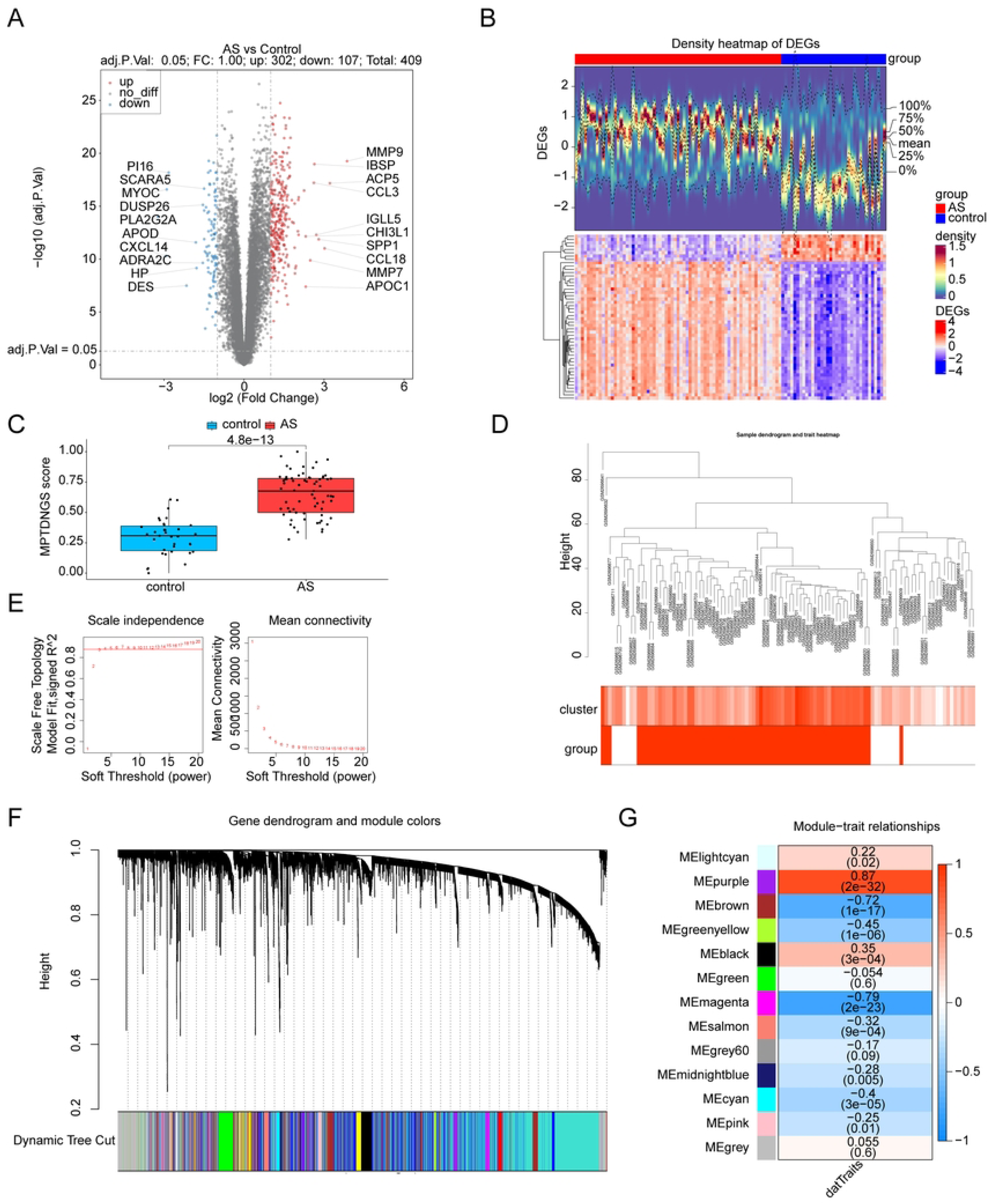
Identification of differentially expressed genes (DEGs) and key module genes. (**A**) Volcanic map of differential genes. (**B**) Heatmap of differential gene expression. (**C**) Mitochondrial permeability transition driven necrosis (MPTDN) score difference between normal sample and atherosclerosis (AS) sample. (**D**) Sample level clustering tree. (**E**) Soft threshold filtering. (**F**) Gene clustering tree. (**G**) Heat map of correlation between modules and phenotypes.

### 3.2 Six DE-MPTDNGs were identified as candidate genes

The key module genes and DEGs were intersected to obtain 313 DE-MPTDNGs **(Figure 2A)**. GO and KEGG analyses of DE-MPTDNGs were performed to explore their possible involvement in biological functions as well as signalling pathways **(Supplementary Table 2-3** Please refer to the appended materials). In terms of GO, they were mainly enrichments of positive regulation of T cell activation and leukocyte mediated immunity, as well as tumor necrosis factor superfamily cytokine production **(Figure 2B)**.The KEGG results showed lipid and atherosclerosis, as well as platelet activation, both of which were pathways clearly associated with AS **(Figure 2C)**. Afterwards, a PPI network was constructed for DE-MPTDNGs to clarify the degree of interactions between them **(Figure 2D)**. Among them, the gene interaction network of MCC Top10 was shown in **Figure 2E**, and it could be seen that integrin, alpha M (ITGAM) was the most important in the network. Eventually, six candidate genes were obtained by selecting genes with MCC Top10 and Degree Top10 to take the intersection for subsequent analyses, namely ITGAM, fc fragment of IgG receptor IIIa (FCGR3A), protein tyrosine phosphatase receptor type C (PTPRC), tumor necrosis factor (TNF), interleukin 1 B (IL1B), and colony stimulating factor 1 receptor (CSF1R) **(Figure 2F)**.

**Figure 2:**
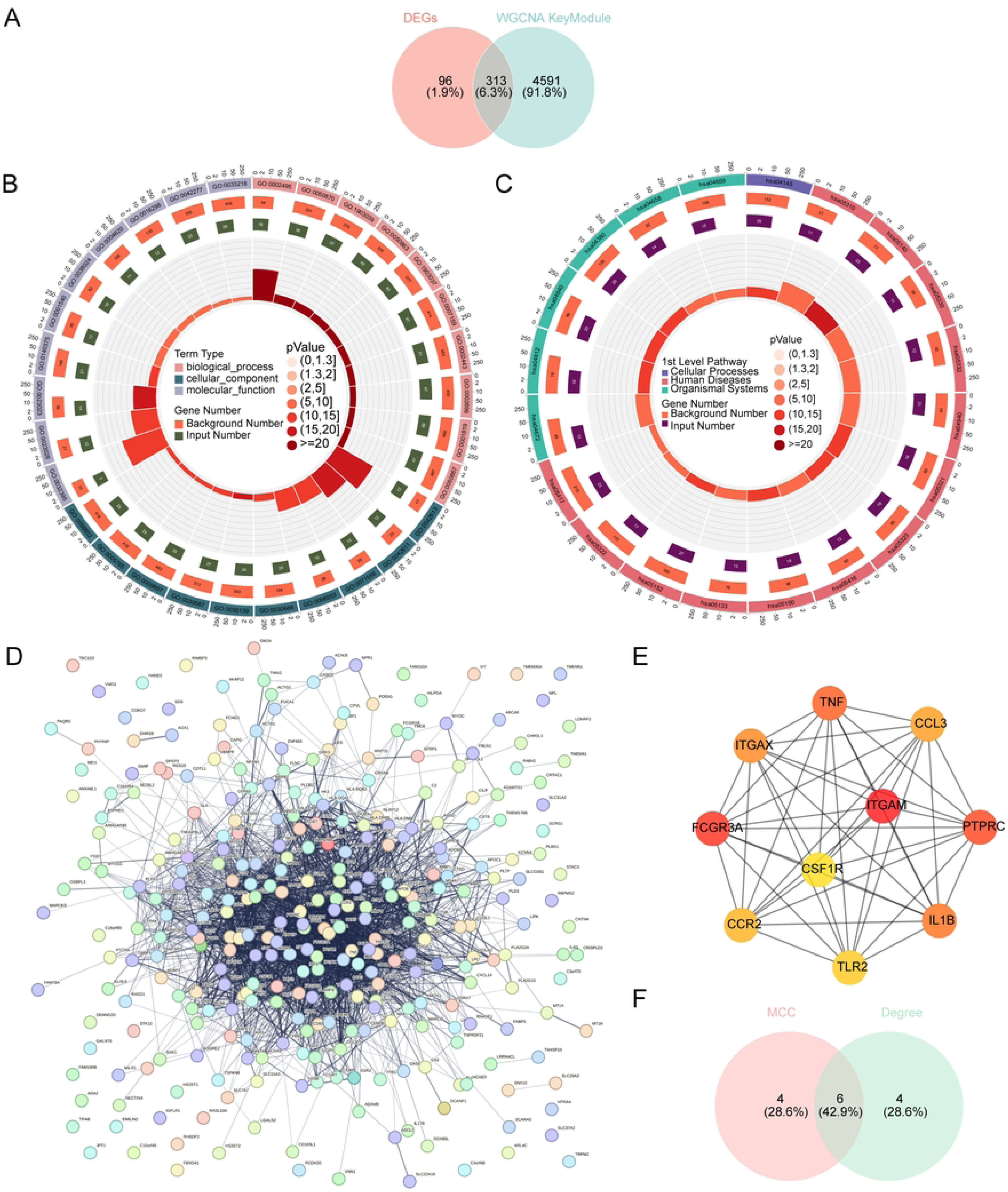
Identification of candidate genes. (**A**) Identification of key differential genes related to MPTDNGS in AS. (**B**) Gene ontology (GO) enrichment circos diagram of key differential genes associated with MPTDNGS. (**C**) Kyoto encyclopedia of genes and genomes (KEGG) enriched circos diagram of key differential genes associated with MPTDNGS. (**D**) PPI protein interaction network of key differential genes associated with MPTDNGS. (**E**) Gene interaction network of MCC Top10. (**F**) Identification of candidate genes.

**Supplementary Table 2**:GO analyses

**Supplementary Table 3**:KEGG analyses

### 3.3 Good ability of biomarkers to differentiate between AS and control samples

LASSO logistic regression analysis **(Figure 3A-B)** and SVM-RFE analysis **(Figure 3C**, **Table 1)** yielded the same five genes **(Figure 3D)**, which were used as feature genes for analysis at a later date. To identify the biomarkers in this study, in the test set GSE100927, the ROC curves revealed that the Area Under Curve (AUC) values of FCGR3A, PTPRC, CSF1R, TNF and ITGAM were exceeded 0.9 **(Figure 4A).** In the validation set GSE66360, since FCGR3A was not present, only the other four genes were analysed, the ROC results showed that the AUC values of all genes were greater than 0.7, except for the gene PTPRC **(Figure 4B)**. These results revealed that most of the feature genes were more powerful for distinguishing AS from control samples. Furthermore, the expression difference results showed that CSF1R, TNF and ITGAM were significantly higher in the AS group than controls, which was in line with the results of the GSE100927 **(Figure 5A-B)**. Therefore, they were defined as biomarkers for this study. To address the biological heterogeneity of disease, we analyzed functional pathways that were active in each of biomarkers. GSEA enrichment results demonsteated that the high level of expression of all three biomarkers mainly regulated natural killer cell mediated cytotoxicity, lysosome, leishmania infection and cytokine cytokine receptor interaction **(Figure 5C)**.

**Figure 3.**
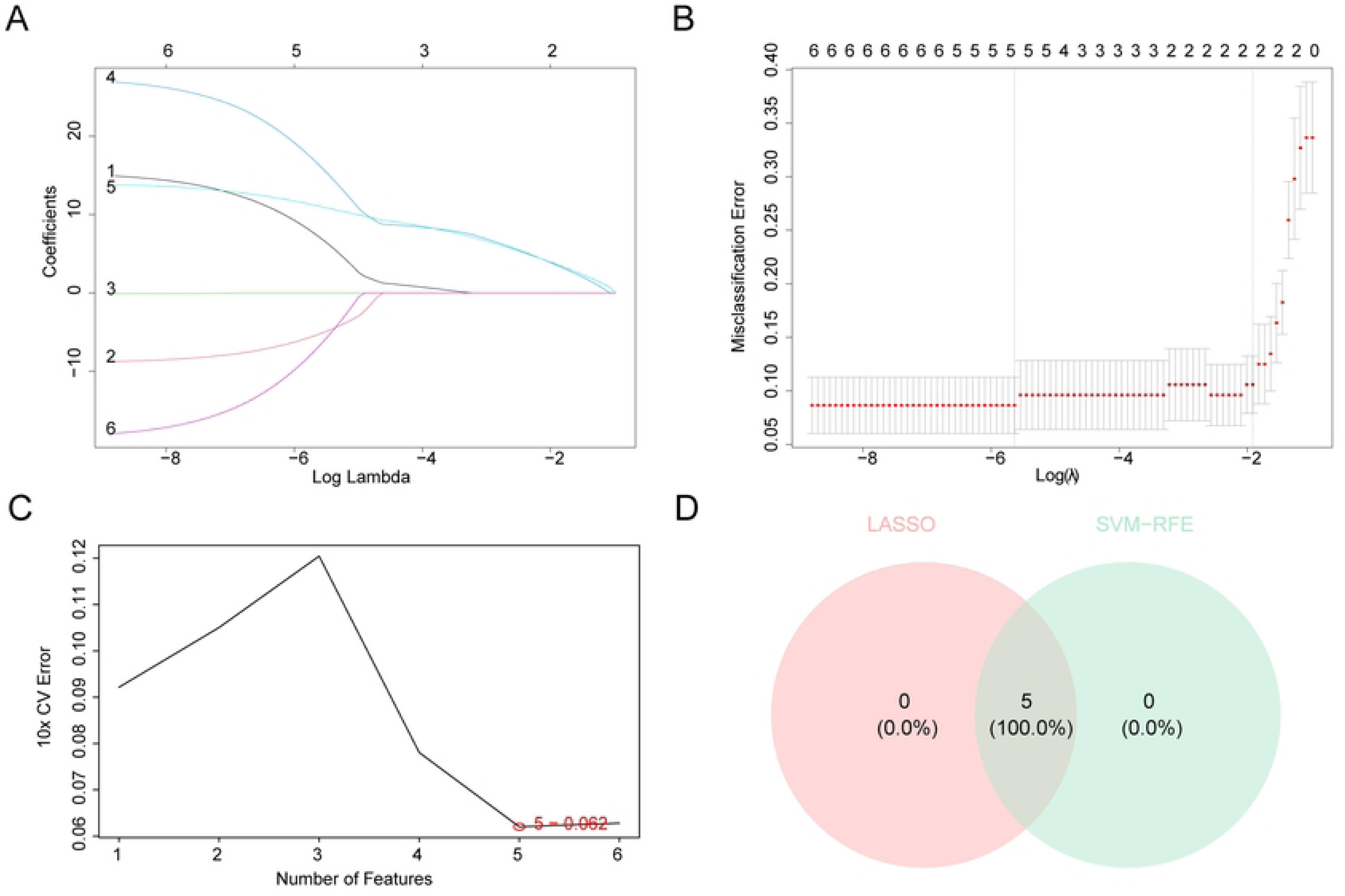
Identifying biomarkers of AS. (**A**) LASSO coefficient spectrum. (**B**) LASSO logic coefficient penalty diagram. (**C**) Support vector machine model error rate. (**D**) Venn diagram of Hub gene.

**Figure 4.**
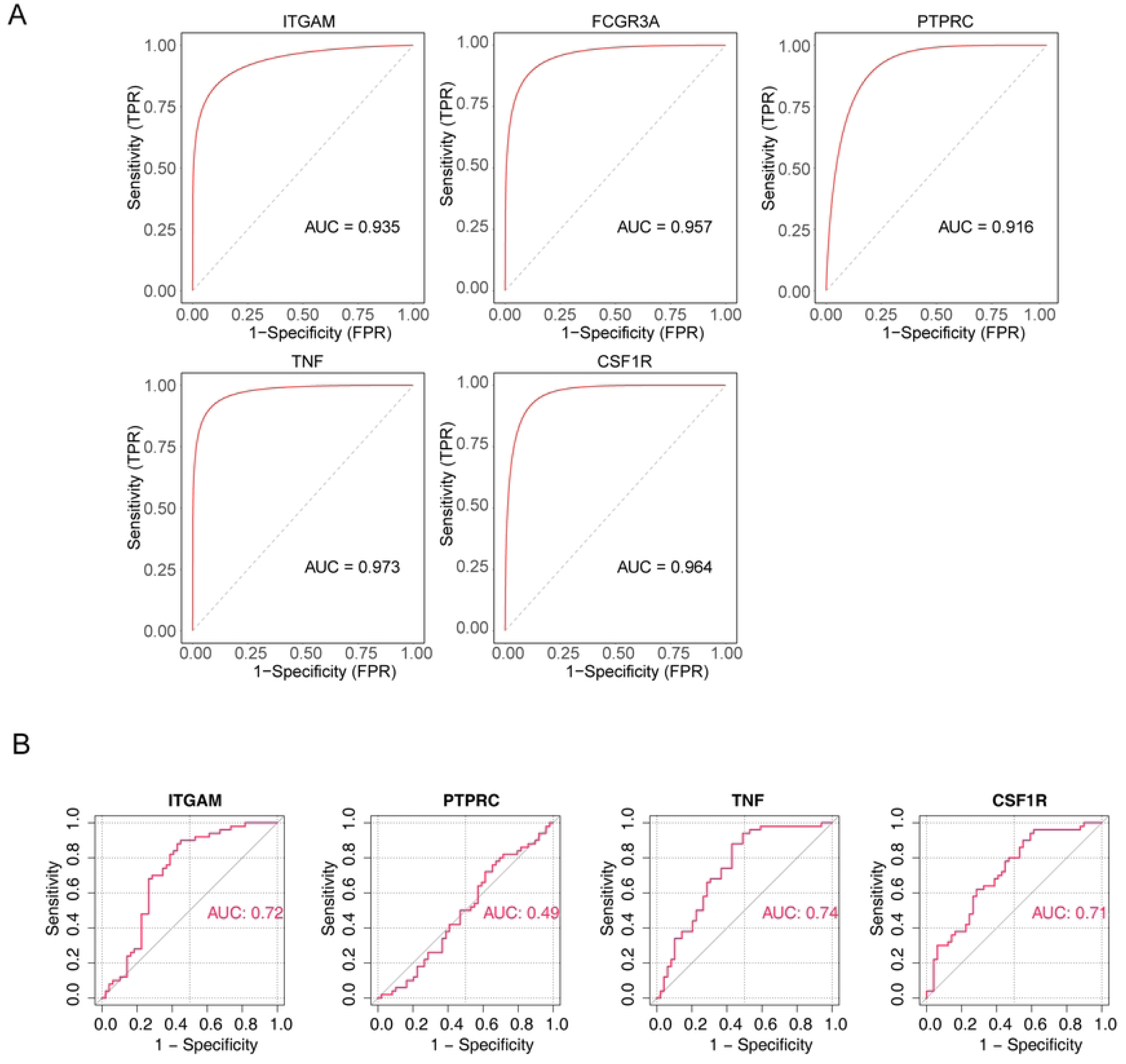
ROC curves of Hub genes in training and validation sets. (**A**) Hub gene ROC curve in training sets (GSE100927). (**B**) Hub gene ROC curve in validation sets (GSE66360).

**Figure 5.**
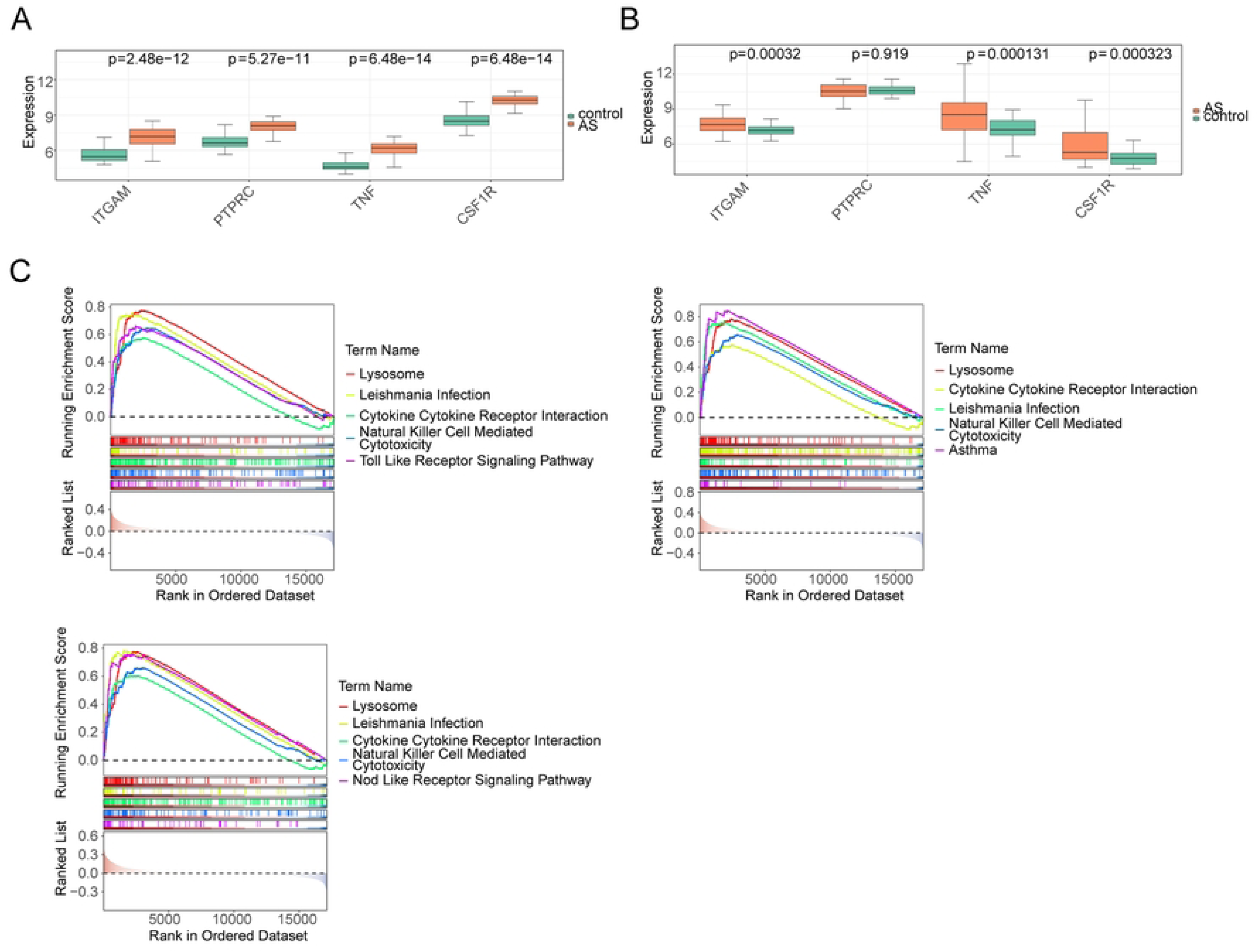
Biomarkers identification and GESA enrichment analysis. (**A-B**) Differences in the expression of Hub gene in training and validation sets. (**C**) GSEA enrichment map of biomarkers.

**Table 1.**
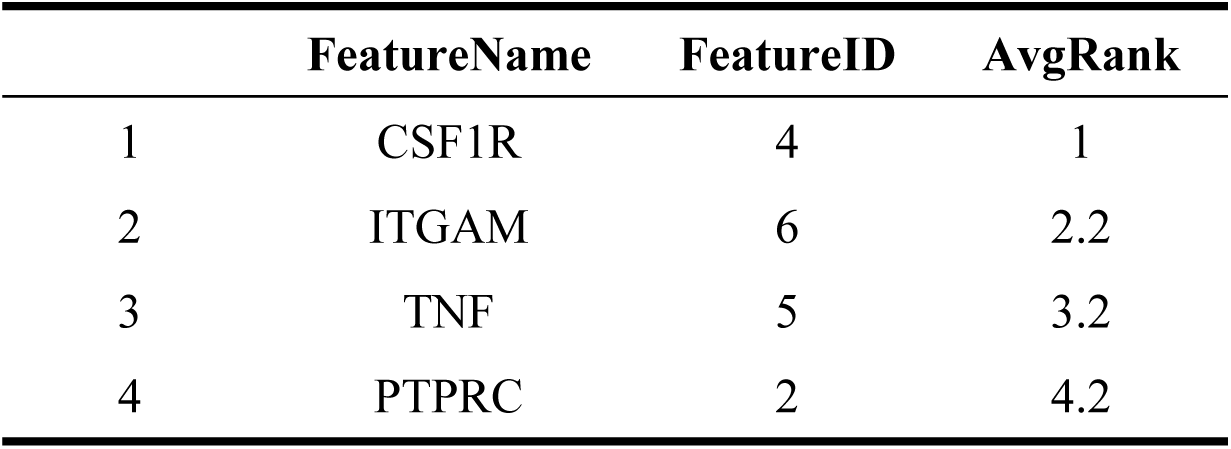
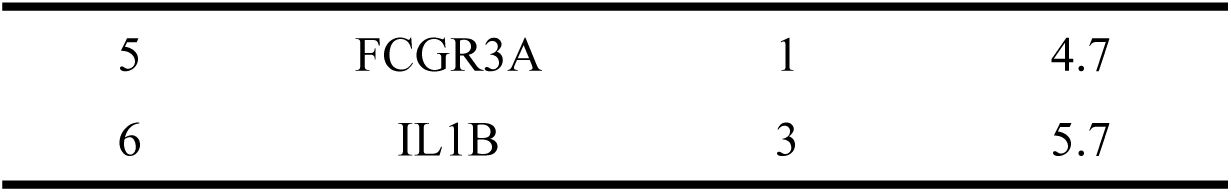
SVM-RFE Model feature gene ranking.

|  | FeatureName | FeatureID | AvgRank |
| --- | --- | --- | --- |
| 1 | CSF1R | 4 | 1 |
| 2 | ITGAM | 6 | 2.2 |
| 3 | TNF | 5 | 3.2 |
| 4 | PTPRC | 2 | 4.2 |
| 5 | FCGR3A | 1 | 4.7 |
| 6 | IL1B | 3 | 5.7 |

### 3.4 ITGAM was of great significance in the immune microenvironment of AS

The clinicopathological significance of immune microenvironment in predicting efficacy and therapeutic effect. The immune infiltration abundance of the remaining 100 samples after screening in the GSE100927 was shown in **Figure 6A**. There were nine different immune cells (regulatory T cells, M0 Macrophages, etc.) between the group of AS and the group of control **(Figure 6B)**. Among them, the strongest negative correlation with CD4 memory resting T cells was found in M0 macrophages, while they had the most significant positive and negative correlation with ITGAM, respectively (|R|= 0.80 & p< 0.001) **(Figure 6C-D)**. Besides, comparing the GSVA scores of the immune response gene sets in the AS group and the control group, it was found that significant differences in all immune response gene sets **(Figure 6E)**. Interestingly, naturalKiller cell cytotoxicity and TGFb family member receptor were positively (R= 0.92) and negatively (R= 0.86) correlated with ITGAM, respectively (p< 0.001) **(Figure 6F)**.

**Figure 6.**
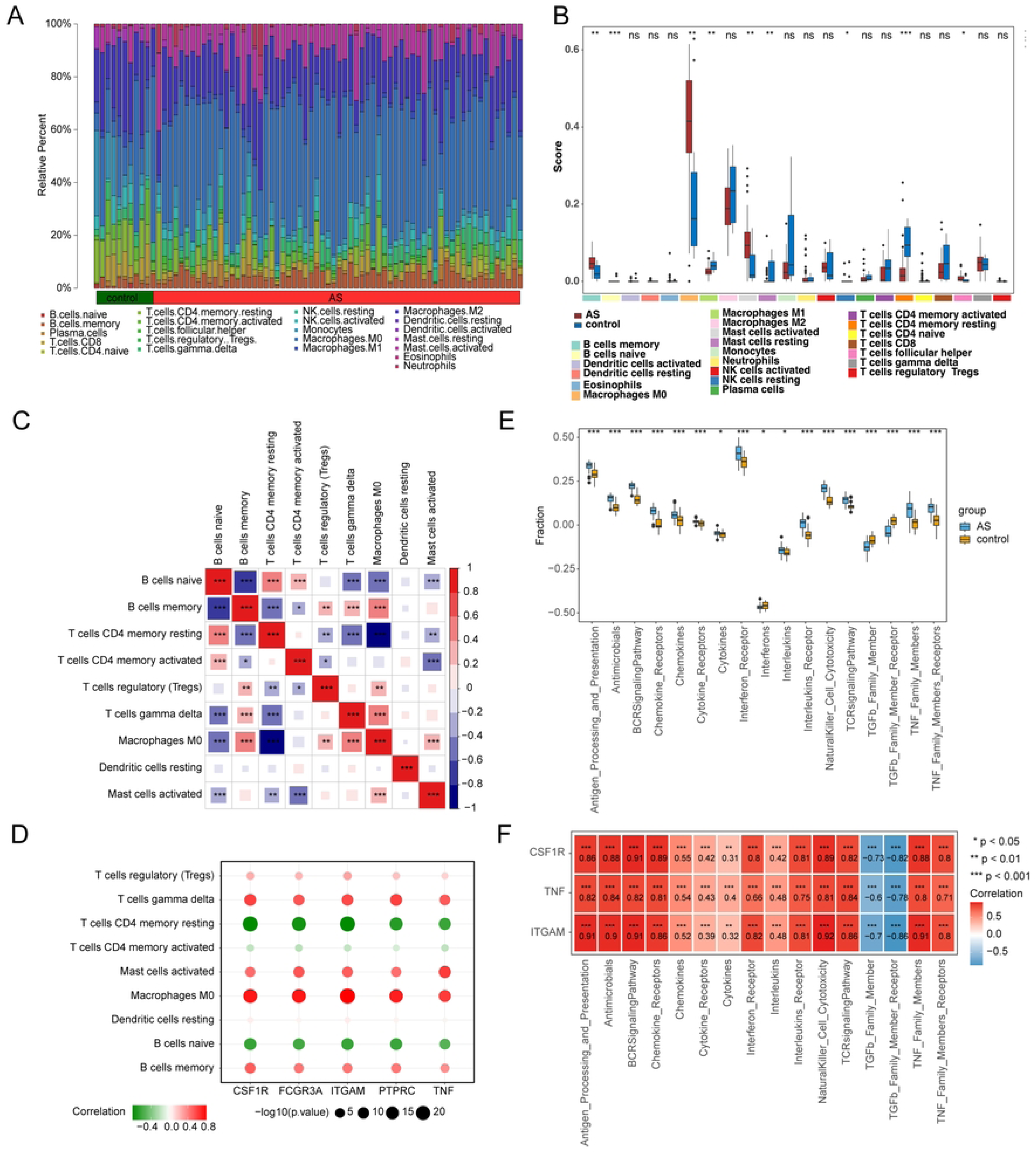
Immune infiltration analysis. (**A**) Abundance of immune cells in AS and control group. (**B**) Map of immune cell infiltration difference between AS and control group. (**C**) Heat map of correlation between differential immune cells. (**D**) Heat map of correlation between Hub gene and differential immune cells. (E) Differences in the activity of immune response gene sets between AS and control groups. (**F**) Correlation analysis between Hub gene and differential immune response gene set.

### 3.5 The molecular regulatory network of biomarkers

A successful ceRNA network was constructed based on 28 miRNAs and 1080 lncRNAs predicted from miRDB and StarBase databases **(Supplementary Figure 1-2)**. For example, hsa-miR-301a-3p, hsa-miR-34c-5p and hsa-miR-342-3p had regulatory effects on TNF, CSF1R and ITGAM, respectively. Similarly, a total of 56 TF-biomarker relationship pairs were predicted based on NetworkAnalyst database **(Supplementary Figure 3)**. Among them, TFAP2A, FOXC1 and NFKB1 could co-interact with CSF1R and TNF, while TFs that could co-interact with TNF and ITGAM to exert regulatory effects included IRF2, MEF2A, TP53, FOXF2 and FOXA1.

### 3.6. Totally 126 biomarker-drug relationship pairs

A total of 126 biomarker-drug relationship pairs were obtained by drug prediction of biomarkers **(Supplementary Figure 4)**. Surprisingly, the common targeted drugs between TNF and CSF1R, ITGAM were SORAFENIB and ATORVASTATIN, respectively. After that, the drugs with the highest Query score and Interaction Score were molecularly docked with biomarkers. The binding energies between CSF1R and ARRY-382, EDICOTINIB were-8.15 kcal/mol and-8.93 kcal/mol, respectively **(Figure 7A-B)**. The corresponding binding energy between TNF and Meropenem was-8.08 kcal/mol **(Figure 7C)**, while ITGAM and LIAROZOLE were-7.12 kcal/mol **(Figure 7D)**. The above results indicate great stability of docking between biomarkers and drugs.

**Figure 7.**
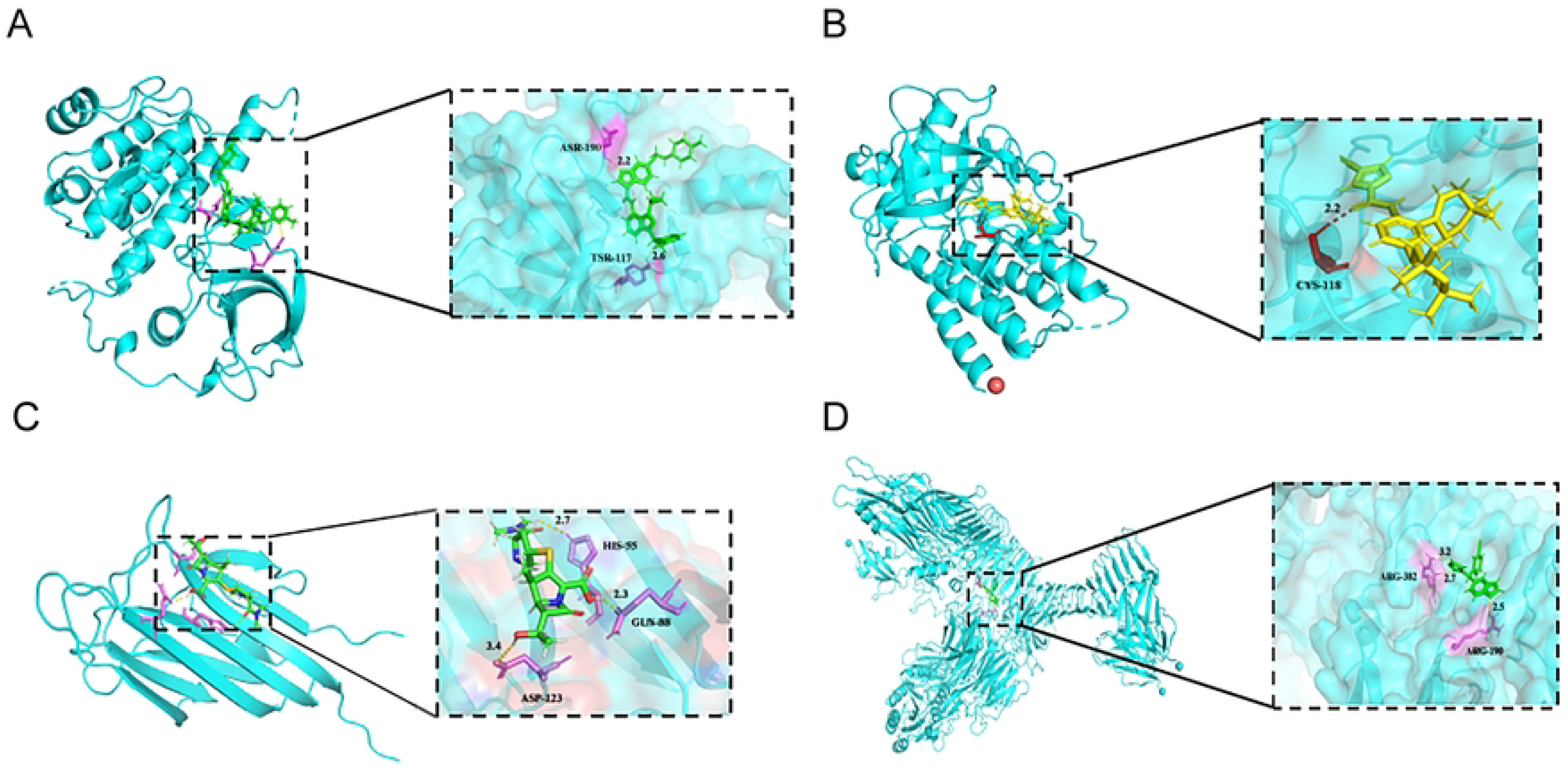
Results of molecular docking. (**A**) Results of molecular docking between CSF1R and ARRY-382. (**B**) CSF1R and EDICOTINIB docking results. (**C**) Results of molecular docking between TNF and Meropenem. (**D**) Results of molecular docking between ITGAM and LIAROZOLE.

**Supplementary Figure 1**:miRNA network

**Supplementary Figure 2**:lncRNA network

**Supplementary Figure 3**:TF network

**Supplementary Figure 4:**Hub gene-drug interaction

### 3.7 Expression validation of biomarkers

We investigated the expression of three biomarkers (CSF1R, TNF and ITGAM) in AS tissues by qRT-PCR and compared it with that of normal adjacent tissues. Our data showed that compared with normal adjacent tissues, the trend of CSF1R expression was consistent with both datasets, being higher in the AS group than in the control group, whereas TNF and ITGAM showed less significant differences between the two groups **(Figure 8)**.

**Figure 8:**
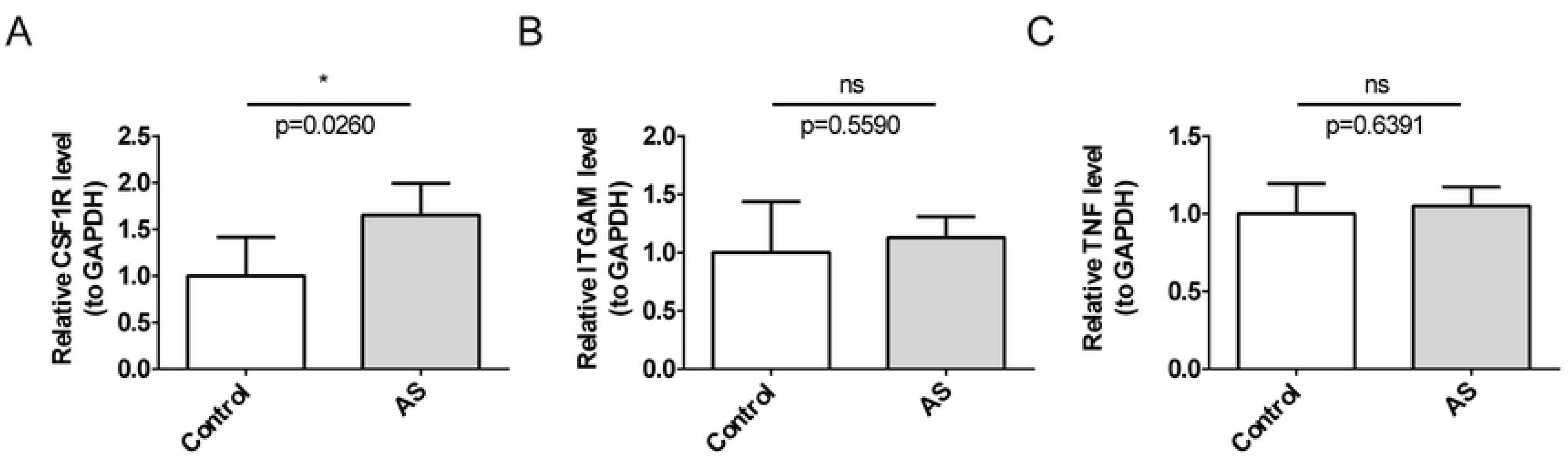
Expression of three biomarkers in AS tissues. (A) CSF1R; (B) ITGAM; (C) TNF.

## 4. Discussion

The aim of this study was to investigate the role of MPTDNGs in AS, a major cause of cardiovascular disease and often associated with it. A comprehensive bioinformatics approach was employed to identify three MPTDN-associated biomarkers within AS: CSF1R, TNF, and ITGAM. We conducted further investigations on these three biomarkers, including GSEA, immune infiltration analysis, construction of the ceRNA regulatory network, drug prediction, and molecular docking. Lastly, qRT-PCR utilized to examine their expression in AS tissues. The results showed that compared to normal adjacent tissues, the trend of CSF1R expression was consistent with both datasets, being higher in the AS group than in the control group, whereas TNF and ITGAM showed less significant differences between the two groups. qRT-PCR and RNA-seq are two fundamentally different technological platforms that employ entirely separate data processing and analysis workflows during their data analysis procedures.qRT-PCR typically uses a relative quantification method to assess gene expression levels by comparing against a standard curve or using an internal reference gene[36]. In contrast, the analysis of RNA-seq data involves several steps including read alignment, expression estimation, and statistical comparison of expression levels across different conditions, with each step potentially introducing variability. Moreover, RNA-seq is capable of detecting low-abundance transcripts that might be difficult to detect with qRT-PCR, leading to discrepancies in the results obtained from the two methods[37].

Colony-stimulating factor 1 receptor (CSF1R) is a member of the Class III receptor tyrosine kinase family. Its primary function of CSF1R is to regulate the homeostasis of macrophages, including their production, differentiation, and function[38, 39]. According to reports, an upregulation of CSF1R expression can expedite the progression of AS. This mechanism behind this involves the colony-stimulating factor 1 (CSF1), secreted by endothelial cells, which regulates the survival, proliferation, and differentiation of macrophages via the CSF1 receptor (CSF1R), thereby promoting the development of AS[1, 40]. Similarly, given the profound impact of CSF1 on the progression of AS, which includes enhancing the production of monocytes, the differentiation of monocytes into macrophages, and the proliferation and survival of macrophages, studies have substantiated the critical role of IL-6 signaling in the development of Tet2-conditional hematopoietic (CH) AS. The primary mechanism for this process also involves the upregulation of CSF1R[41].

Tumor necrosis factor (TNF) is a cytokine primarily produced by activated macrophages. Its main functions include mediating inflammatory responses and anti-tumor activities, such as tumor cell apoptosis, inhibiting tumor growth and metastasis, and enhancing the killing effect of immune cells on tumor cells[42, 43]. Research has shown that the TNF gene can promote endothelial inflammation and advance the process of AS. The pro-inflammatory and pro-atherosclerotic signals mediated by TNF primarily originate from TNFR1. This receptor, through a ubiquitin E3 ligase pathway distinct from TRAF6, involving TRAF2 or TRAF5, cIAP1 and cIAP2, as well as the ubiquitin ligase complex LUBAC, triggers the canonical activation of NFκB, thereby exacerbating endothelial inflammation and the progression of AS[44, 45]. In the realm of liver disease research, Etsuro Hatano discovered that using the MPT inhibitors Cyclosporin A and Trifluoperazine in combination can protect liver cells infected with adenovirus-adIKKappabrs from apoptosis mediated by TNF[46]. Ethanol exposure enhances TNF-α-induced cell death in cells, which is dependent on the development of MPT[47].

ITGAM, also known as CD11b, is a membrane protein expressed on the surface of leukocytes and belongs to the Immunoglobulin Superfamily (IgSF)[48]. ITGAM is associated with cell adhesion, chemotaxis, and migration. It mediates macrophage adhesion and transendothelial migration, leading to macrophage infiltration and arterial inflammation[49]. Research indicates that the progression of AS is associated with the ITGAM gene, which may interact with immune cells through some as yet unidentified cell membrane receptors or ligands[50]. Sentong Xiao and Chunyan Kuang discovered that ITGAM is involved in the pathogenesis of acute myocardial infarction (AMI)[51]. To date, there have been scant reports on the association of these three genes with MPTDN in research, indicating that our study is the first to identify the differential expression of these three biomarkers in AS as regulated by an MPT-driven necrotic pathway.

GSEA revealed that the TNF, ITGAM, and CSF1R genes were primarily enriched in pathways associated with the lysosome, Leishmania infection, cytokine-cytokine receptor interaction, and natural killer cell mediated cytotoxicity. The lysosome serves as a pivotal degradation center and signaling hub that plays a crucial role in cellular homeostasis, development, and the aging process[52]. Studies suggest that macrophage lysosomes are implicated in virtually the entire progression of AS. Dysfunction of lysosomal activity may lead to pathological alterations by modulating autophagy, inflammasomes, apoptosis, and lysosomal biogenesis, thereby contributing to the development of AS[53]. Infections with Leishmania parasites trigger macrophage activation, intensifying the inflammatory component of AS and accelerating its onset[54]. Cytokine receptor interactions also play a pivotal role in the pathogenesis of AS. For example, IFN-γ communicates via a heterodimeric cell surface receptor composed of two distinct subunits, promoting the activation of endothelial cells and the mobilization of leukocytes, as well as exacerbating inflammation (by upregulating IL-1β, IL-6, and CCL5 and downregulating IL-1Ra). This shifts macrophages toward an atherosclerotic M1 profile, polarizes T cells towards a Th1 phenotype, inhibits collagen synthesis, and ultimately results in plaque instability[55]. Reports indicate that in elderly individuals, a reduction in the quantity and cytotoxicity of natural killer cells is associated with AS and coronary artery disease, mechanisms of which are linked to inflammatory activity[56, 57]. In sum, the biological processes we have identified as enriched suggest that these biomarkers may regulate AS via modulating macrophages and inflammatory activity.

According to the analysis of correlations between genes and immune cells, there was a notably strong positive relationship between M0 Macrophages and ITGAM, while a profoundly negative correlation exists between CD4 memory resting T cells and ITGAM. Bin Wang and Dan Zhao propose that the progression of AS may be associated with ITGAM, which appears to regulate immune-competent cells such as M0 Macrophages. The underlying mechanism might involve the gene interacting with immune cells through some yet unidentified cell membrane receptors or ligands[50]. Research has indicated that hyperbaric oxygen treatment affects ITGAM expression and the infiltration of CD4 lymphocyte T cells, subsequently influencing scar keloid formation. This may also be a mechanism by which AS is affected[58]. Intriguingly, within the domain of immune gene set analysis, TGF-β family member receptor also exhibited a markedly negative correlation with ITGAM. Beta (TGF-β) plays a key role in the proliferation, differentiation, migration, tube formation, and sprouting of endothelial cells and smooth muscle cells. Dysregulation of TGF-β signaling in the human body can lead to vascular pathologies and cardiovascular diseases, such as arteriovenous malformations, aneurysms, and atherosclerosis[59].TGF-β exerts its biological effects through specific membrane-bound receptors, such as Type I and Type II TGF-β receptors, acting as upregulators or downregulators. It is reported that FOXP3 positively regulates the production of abundant anti-inflammatory mediators by macrophages, including Interleukin 10 (IL-10), Transforming Growth Factor Beta (TGF-β), and Arginase 1 (ARG1). These anti-inflammatory mediators contribute to modulating the inflammatory response, promoting tissue repair, and limiting excessive inflammation, thus highlighting the role of FOXP3 in the resolution of inflammation through macrophages. FOXP3 exhibits a predilection for resolving inflammation, as evidenced by its regulation of anti-inflammatory mediator production and direct upregulation of the anti-inflammatory gene ITGAM, playing a pivotal role in modulating immune response and maintaining tissue homeostasis[60]. The progression of AS is closely related to inflammatory responses, thereby suggesting that ITGAM may also mediate atherosclerotic effects by modulating inflammation through this pathway.

This analysis synthesizes various methodologies, with the primary result being the identification of biomarkers driving necrosis in AS, mediated by MPT, and these findings have been substantiated quite convincingly. These biomarkers represent a solid foundation and direction for future research endeavors.Although this research has achieved certain accomplishments, it inevitably confronts several limitations. Firstly, owing to the sample size of the dataset, the biomarkers identified in this study might not have met the extensive criteria for applicability. To promote its clinical application, there is an urgent need for larger datasets to offer more robust support. For this purpose, we intend to actively seek cooperation opportunities with other institutions in the future to expand the sample size and further enhance the reliability and universality of our findings through cross-validation of multiple datasets. Secondly, this study mainly relies on qRT-PCR technology during the verification process, and there is still a dearth of evidence from other experimental approaches. To verify our findings more comprehensively and profoundly, we plan to conduct a variety of experiments, including Western blotting, gene knockout or enhancement, and animal models, etc., to form a more complete verification system, thereby strengthening the persuasiveness of the research conclusions. Finally, we acknowledge that the current research is still at the preliminary stage, and further exploration of the involved mechanisms remains pending. Therefore, in future work, we will continue to deepen the research content and refine the research direction, aiming to reveal more detailed and in-depth biological mechanisms and contribute more scientific strength to the development of related fields.

## List of abbreviations

Abbreviations: Full name in English
AS: Atherosclerosis
MPTDN: Mitochondrial permeability transition driven necrosis
MPTDNGs: MPTDN-associated genes
WGCNA: Weighted correlation network analysis
DEGs: Differentially expressed genes
RCC: Renal cell carcinoma
DE-MPTDNGs: Differentially expressed MPTDNGs PPI Protein-protein interaction
GO: Gene ontology
CSF1R: Colony stimulating factor 1 receptor
KEGG: Kyoto encyclopedia of genes and genomes
AUC: Area under curve
ROC: Receiver operating characteristic
GSEA: Gene set enrichment analysis
ssGSEA: Single-sample gene set enrichment analysis
GSVA: Gene set variation analysis
TNF: Tumor necrosis factor
ITGAM: Integrin, alpha M
MPT: Mitochondrial permeability transition
MMC: Maximal clique centrality
TGFb: Transforming growth factor-b
LASSO: Least absolute shrinkage and selection operator
SVM-RFE: Support vector machines-recursive feature elimination

## Data Availability

The data that support the findings of this study are openly available in Gene Expression Omnibus (GEO) at https://www.ncbi.nlm.nih.gov/geo/, reference number GSE100927?GSE66360.

## Acknowledgements

Not applicable.

## Availability of data and materials

The data that support the findings of this study are openly available in Gene Expression Omnibus (GEO) at https://www.ncbi.nlm.nih.gov/geo/, reference number GSE100927、GSE66360.

## Authors’ contributions

YHJ, YXS and FN confirm the authenticity of all the raw data. YHJ, YXS and FN were responsible for the concep-tualization, methodology and design of the research, as well as writing and preparing the original draft. SYW, YL, were responsible for the bioinformatic data collection and analysis.SYW, YL, JWZ were responsible for the experimental data acquisition and analysis. YL, JWZ were responsible for the software validation and result interpretation.JWZ was responsible for the figure preparation. All authors have read and approved the final manuscript.

## Ethics approval and consent to participate

The present study was approved by the Affiliated Hospital of Shandong University of Traditional Chinese Medicine (Jinan, China; approval no. 2023-71-KY; July 11, 2023) and complied with The Declaration of Helsinki. Written informed consent was obtained from all subjects.

## Patient consent for publication

Not applicable.

## Supplementary Figure/Table legends

Supplementary Figure 1 miRNA network

Supplementary Figure 2 lncRNA network

Supplementary Figure 3 TF network Supplementary

Figure 4 Hub gene-drug interaction

Supplementary Table 1 Sequence information of primers

Supplementary Table 2 GO analyses

Supplementary Table 3 KEGG analyses

## References

1. Libby P. The changing landscape of atherosclerosis. Nature. (2021) 592(7855):524-33. doi: 10.1038/s41586-021-03392-8

2. Libby P, Hansson GK. From Focal Lipid Storage to Systemic Inflammation: JACC Review Topic of the Week. J Am Coll Cardiol. (2019) 74(12):1594–607. doi: 10.1016/j.jacc.2019.07.061

3. Becattini C, Dentali F, Camporese G, Sembolini A, Rancan E, Tonello C, et al. Carotid atherosclerosis and risk for ischemic stroke in patients with atrial fibrillation on oral anticoagulant treatment. Atherosclerosis. (2018) 271:177–81. doi: 10.1016/j.atherosclerosis.2018.02.004

4. Tuttolomondo A, Di Raimondo D, Pecoraro R, Arnao V, Pinto A, Licata G. Atherosclerosis as an inflammatory disease. Curr Pharm Des. (2012) 18(28):4266–88. doi: 10.2174/138161212802481237

5. Shaw LJ, Blankstein R, Min JK. Outcomes in Stable Coronary Disease: Is Defining High-Risk Atherosclerotic Plaque Important. J Am Coll Cardiol. (2019) 73(3):302–4. doi: 10.1016/j.jacc.2018.11.017

6. Wong MC, Zhang DX, Wang HH. Rapid emergence of atherosclerosis in Asia: a systematic review of coronary atherosclerotic heart disease epidemiology and implications for prevention and control strategies. Curr Opin Lipidol. (2015) 26(4):257–69. doi: 10.1097/MOL.0000000000000191

7. Herrington W, Lacey B, Sherliker P, Armitage J, Lewington S. Epidemiology of Atherosclerosis and the Potential to Reduce the Global Burden of Atherothrombotic Disease. Circ Res. (2016) 118(4):535–46. doi: 10.1161/CIRCRESAHA.115.307611

8. Arnett DK, Blumenthal RS, Albert MA, Buroker AB, Goldberger ZD, Hahn EJ, et al. 2019 ACC/AHA Guideline on the Primary Prevention of Cardiovascular Disease: A Report of the American College of Cardiology/American Heart Association Task Force on Clinical Practice Guidelines. Circulation. (2019) 140(11):e596-596e646. doi: 10.1161/CIR.0000000000000678

9. Knuuti J, Wijns W, Saraste A, Capodanno D, Barbato E, Funck-Brentano C, et al. 2019 ESC Guidelines for the diagnosis and management of chronic coronary syndromes. Eur Heart J. (2020) 41(3):407-77. doi: 10.1093/eurheartj/ehz425

10. Bułdak Ł. Cardiovascular Diseases-A Focus on Atherosclerosis, Its Prophylaxis, Complications and Recent Advancements in Therapies. Int J Mol Sci. (2022) 23(9):4695. doi: 10.3390/ijms23094695

11. Izzo V, Bravo-San Pedro JM, Sica V, Kroemer G, Galluzzi L. Mitochondrial Permeability Transition: New Findings and Persisting Uncertainties. Trends Cell Biol. (2016) 26(9):655–67. doi: 10.1016/j.tcb.2016.04.006

12. Vanden Berghe T, Linkermann A, Jouan-Lanhouet S, Walczak H, Vandenabeele P. Regulated necrosis: the expanding network of non-apoptotic cell death pathways. Nat Rev Mol Cell Biol. (2014) 15(2):135–47. doi: 10.1038/nrm3737

13. Liu J, Zhang M, Sun Q, Qin X, Gao T, Xu Y, et al. Construction of a novel MPT-driven necrosis-related lncRNAs signature for prognosis prediction in laryngeal squamous cell carcinoma. Environ Sci Pollut Res Int. (2023) 30(31):77210–25. doi: 10.1007/s11356-023-26996-1

14. Dhaouadi N, Vitto V, Pinton P, Galluzzi L, Marchi S. Ca(2+) signaling and cell death. Cell Calcium. (2023) 113:102759. doi: 10.1016/j.ceca.2023.102759

15. Lemasters JJ, Qian T, Elmore SP, Trost LC, Nishimura Y, Herman B, et al. Confocal microscopy of the mitochondrial permeability transition in necrotic cell killing, apoptosis and autophagy. Biofactors. (1998) 8(3-4):283–5. doi: 10.1002/biof.5520080316

16. Reznik E, Luna A, Aksoy BA, Liu EM, La K, Ostrovnaya I, et al. A Landscape of Metabolic Variation across Tumor Types. Cell Syst. (2018) 6(3):301–13.e3. doi: 10.1016/j.cels.2017.12.014

17. Lemasters JJ, Qian T, He L, Kim JS, Elmore SP, Cascio WE, et al. Role of mitochondrial inner membrane permeabilization in necrotic cell death, apoptosis, and autophagy. Antioxid Redox Signal. (2002) 4(5):769–81. doi: 10.1089/152308602760598918

18. Naumova N, Šachl R. Regulation of Cell Death by Mitochondrial Transport Systems of Calcium and Bcl-2 Proteins. Membranes (Basel). (2020) 10(10):299. doi: 10.3390/membranes10100299

19. Uribe P, Cabrillana ME, Fornés MW, Treulen F, Boguen R, Isachenko V, et al. Nitrosative stress in human spermatozoa causes cell death characterized by induction of mitochondrial permeability transition-driven necrosis. Asian J Androl. (2018) 20(6):600–7. doi: 10.4103/aja.aja_29_18

20. Robichaux DJ, Harata M, Murphy E, Karch J. Mitochondrial permeability transition pore-dependent necrosis. J Mol Cell Cardiol. (2023) 174:47–55. doi: 10.1016/j.yjmcc.2022.11.003

21. Bauer TM, Murphy E. Role of Mitochondrial Calcium and the Permeability Transition Pore in Regulating Cell Death. Circ Res. (2020) 126(2):280–93. doi: 10.1161/CIRCRESAHA.119.316306

22. Ritchie ME, Phipson B, Wu D, Hu Y, Law CW, Shi W, et al. limma powers differential expression analyses for RNA-sequencing and microarray studies. Nucleic Acids Res. (2015) 43(7):e47. doi: 10.1093/nar/gkv007

23. Liang K, Guo Z, Zhang S, Chen D, Zou R, Weng Y, et al. GPR37 expression as a prognostic marker in gliomas: a bioinformatics-based analysis. Aging (Albany NY). (2023) 15(19):10146–67. doi: 10.18632/aging.205063

24. Gu Z, Eils R, Schlesner M. Complex heatmaps reveal patterns and correlations in multidimensional genomic data. Bioinformatics. (2016) 32(18):2847–9. doi: 10.1093/bioinformatics/btw313

25. Hänzelmann S, Castelo R, Guinney J. GSVA: gene set variation analysis for microarray and RNA-seq data. BMC Bioinformatics. (2013) 14:7. doi: 10.1186/1471-2105-14-7

26. Langfelder P, Horvath S. WGCNA: an R package for weighted correlation network analysis. BMC Bioinformatics. (2008) 9:559. doi: 10.1186/1471-2105-9-559

27. Wu T, Hu E, Xu S, Chen M, Guo P, Dai Z, et al. clusterProfiler 4.0: A universal enrichment tool for interpreting omics data. Innovation (Camb). (2021) 2(3):100141. doi: 10.1016/j.xinn.2021.100141

28. Shannon P, Markiel A, Ozier O, Baliga NS, Wang JT, Ramage D, et al. Cytoscape: a software environment for integrated models of biomolecular interaction networks. Genome Res. (2003) 13(11):2498–504. doi: 10.1101/gr.1239303

29. Simon N, Friedman J, Hastie T, Tibshirani R. Regularization Paths for Cox’s Proportional Hazards Model via Coordinate Descent. J Stat Softw. (2011) 39(5):1–13. doi: 10.18637/jss.v039.i05

30. Asim Shahid M, Alam MM, Mohd Su’ud M. Improved accuracy and less fault prediction errors via modified sequential minimal optimization algorithm. PLoS One. (2023) 18(4):e0284209. doi: 10.1371/journal.pone.0284209

31. Kuleshov MV, Jones MR, Rouillard AD, Fernandez NF, Duan Q, Wang Z, et al. Enrichr: a comprehensive gene set enrichment analysis web server 2016 update. Nucleic Acids Res. (2016) 44(W1):W90–7. doi: 10.1093/nar/gkw377

32. Shurson GC, Ku PK, Waxler GL, Yokoyama MT, Miller ER. Physiological relationships between microbiological status and dietary copper levels in the pig. J Anim Sci. (1990) 68(4):1061–71. doi: 10.2527/1990.6841061x

33. N’dilimabaka N, Mounguegui DM, Lekana-Douki SE, Yattara MK, Obame-Nkoghe J, Longo-Pendy NM, et al. Biochemical and hematological factors associated with COVID-19 severity among Gabonese patients: A retrospective cohort study. Front Cell Infect Microbiol. (2022) 12:975712. doi: 10.3389/fcimb.2022.975712

34. Guo J, Mei ZW, Wang XJ, Li Q, Qin J. Molecular docking and network pharmacological analysis of Scutellaria baicalensis against renal cell carcinoma. Eur Rev Med Pharmacol Sci. (2023) 27(23):11574–86. doi: 10.26355/eurrev_202312_34596

35. Livak KJ, Schmittgen TD. Analysis of relative gene expression data using real-time quantitative PCR and the 2(-Delta Delta C(T)) Method. Methods. (2001) 25(4):402–8. doi: 10.1006/meth.2001.1262

36. Jozefczuk J, Adjaye J. Quantitative real-time PCR-based analysis of gene expression. Methods Enzymol. (2011) 500:99–109. doi: 10.1016/B978-0-12-385118-5.00006-2

37. Chung M, Bruno VM, Rasko DA, Cuomo CA, Muñoz JF, Livny J, et al. Best practices on the differential expression analysis of multi-species RNA-seq. Genome Biol. (2021) 22(1):121. doi: 10.1186/s13059-021-02337-8

38. Benner B, Good L, Quiroga D, Schultz TE, Kassem M, Carson WE, et al. Pexidartinib, a Novel Small Molecule CSF-1R Inhibitor in Use for Tenosynovial Giant Cell Tumor: A Systematic Review of Pre-Clinical and Clinical Development. Drug Des Devel Ther. (2020) 14:1693–704. doi: 10.2147/DDDT.S253232

39. Mo H, Hao Y, Lv Y, Chen Z, Shen J, Zhou S, et al. Overexpression of macrophage-colony stimulating factor-1 receptor as a prognostic factor for survival in cancer: A systematic review and meta-analysis. Medicine (Baltimore). (2021) 100(12):e25218. doi: 10.1097/MD.0000000000025218

40. Wei Y, Zhu M, Corbalán-Campos J, Heyll K, Weber C, Schober A. Regulation of Csf1r and Bcl6 in macrophages mediates the stage-specific effects of microRNA-155 on atherosclerosis. Arterioscler Thromb Vasc Biol. (2015) 35(4):796–803. doi: 10.1161/ATVBAHA.114.304723

41. Liu W, Yalcinkaya M, Maestre IF, Olszewska M, Ampomah PB, Heimlich JB, et al. Blockade of IL-6 signaling alleviates atherosclerosis in Tet2-deficient clonal hematopoiesis. Nat Cardiovasc Res. (2023) 2(6):572–86. doi: 10.1038/s44161-023-00281-3

42. Xue Y, Zeng X, Tu WJ, Zhao J. Tumor Necrosis Factor-α: The Next Marker of Stroke. Dis Markers. (2022) 2022:2395269. doi: 10.1155/2022/2395269

43. Holbrook J, Lara-Reyna S, Jarosz-Griffiths H, McDermott M. Tumour necrosis factor signalling in health and disease. F1000Res. (2019) 8:F1000 Faculty Rev-111 [pii]. doi: 10.12688/f1000research.17023.1

44. Gao W, Liu H, Yuan J, Wu C, Huang D, Ma Y, et al. Exosomes derived from mature dendritic cells increase endothelial inflammation and atherosclerosis via membrane TNF-α mediated NF-κB pathway. J Cell Mol Med. (2016) 20(12):2318–27. doi: 10.1111/jcmm.12923

45. Jean-Charles PY, Wu JH, Zhang L, Kaur S, Nepliouev I, Stiber JA, et al. USP20 (Ubiquitin-Specific Protease 20) Inhibits TNF (Tumor Necrosis Factor)-Triggered Smooth Muscle Cell Inflammation and Attenuates Atherosclerosis. Arterioscler Thromb Vasc Biol. (2018) 38(10):2295–305. doi: 10.1161/ATVBAHA.118.311071

46. Hatano E. Tumor necrosis factor signaling in hepatocyte apoptosis. J Gastroenterol Hepatol. (2007) 22 Suppl 1:S43–4. doi: 10.1111/j.1440-1746.2006.04645.x

47. Pastorino JG, Shulga N, Hoek JB. TNF-alpha-induced cell death in ethanol-exposed cells depends on p38 MAPK signaling but is independent of Bid and caspase-8. Am J Physiol Gastrointest Liver Physiol. (2003) 285(3):G503–16. doi: 10.1152/ajpgi.00442.2002

48. Zhang YL, Bai J, Yu WJ, Lin QY, Li HH. CD11b mediates hypertensive cardiac remodeling by regulating macrophage infiltration and polarization. J Adv Res. (2024) 55:17–31. doi: 10.1016/j.jare.2023.02.010

49. Lou Y, Li PH, Liu XQ, Wang TX, Liu YL, Chen CC, et al. ITGAM-mediated macrophages contribute to basement membrane damage in diabetic nephropathy and atherosclerosis. BMC Nephrol. (2024) 25(1):72. doi: 10.1186/s12882-024-03505-1

50. Zhao B, Wang D, Liu Y, Zhang X, Wan Z, Wang J, et al. Six-Gene Signature Associated with Immune Cells in the Progression of Atherosclerosis Discovered by Comprehensive Bioinformatics Analyses. Cardiovasc Ther. (2020) 2020:1230513. doi: 10.1155/2020/1230513

51. Xiao S, Kuang C. Identification of crucial genes that induce coronary atherosclerosis through endothelial cell dysfunction in AMI-identifying hub genes by WGCNA. Am J Transl Res. (2022) 14(11):8166–74.

52. Yang C, Wang X. Lysosome biogenesis: Regulation and functions. J Cell Biol. (2021) 220(6):e202102001. doi: 10.1083/jcb.202102001

53. Zhang Z, Yue P, Lu T, Wang Y, Wei Y, Wei X. Role of lysosomes in physiological activities, diseases, and therapy. J Hematol Oncol. (2021) 14(1):79. doi: 10.1186/s13045-021-01087-1

54. Fernandes LR, Ribeiro AC, Segatto M, Santos LF, Amaral J, Portugal LR, et al. Leishmania major Self-Limited Infection Increases Blood Cholesterol and Promotes Atherosclerosis Development. Cholesterol. (2013) 2013:754580. doi: 10.1155/2013/754580

55. Tsioufis P, Theofilis P, Tsioufis K, Tousoulis D. The Impact of Cytokines in Coronary Atherosclerotic Plaque: Current Therapeutic Approaches. Int J Mol Sci. (2022) 23(24):15937. doi: 10.3390/ijms232415937

56. Bruunsgaard H, Pedersen AN, Schroll M, Skinhøj P, Pedersen BK. Decreased natural killer cell activity is associated with atherosclerosis in elderly humans. Exp Gerontol. (2001) 37(1):127–36. doi: 10.1016/s0531-5565(01)00162-0

57. Jonasson L, Backteman K, Ernerudh J. Loss of natural killer cell activity in patients with coronary artery disease. Atherosclerosis. (2005) 183(2):316–21. doi: 10.1016/j.atherosclerosis.2005.03.011

58. Wang CH, Shan MJ, Liu H, Hao Y, Song KX, Wu HW, et al. Hyperbaric oxygen treatment on keloid tumor immune gene expression. Chin Med J (Engl). (2021) 134(18):2205–13. doi: 10.1097/CM9.0000000000001780

59. Goumans MJ, Ten Dijke P. TGF-β Signaling in Control of Cardiovascular Function. Cold Spring Harb Perspect Biol. (2018) 10(2):a022210. doi: 10.1101/cshperspect.a022210

60. Cai W, Hu M, Li C, Wu R, Lu D, Xie C, et al. FOXP3+ macrophage represses acute ischemic stroke-induced neural inflammation. Autophagy. (2023) 19(4):1144–63. doi: 10.1080/15548627.2022.2116833

